# Schistosomiasis mansoni-alcoholism comorbidity: prevalence and risk factors among adults in Makenene, Cameroon

**DOI:** 10.1101/2025.10.28.25338939

**Authors:** Emmanuelle S. Yimgoua, Christian M. Kenfack, Nestor G. Feussom, Joseph B. Fassi-Kadji, Emilienne T. Nkondo, Ulrich M. Femoe, Louis-Albert Tchuem Tchuente, Hermine B. Jatsa

## Abstract

**Background:** Alcohol consumption in Cameroon is approximately twice the African average, and Makenene is a hotspot of Schistosoma mansoni infection in the country. Considering that both schistosomiasis mansoni and alcoholism induced liver pathology and are public health concerns, this study aimed to investigate the prevalence and risk factors of S. mansoni and alcoholism comorbidity in Makenene, Cameroon.

**Methodology/Principal findings:** A cross-sectional survey was conducted in Makenene, including 431 adults from four neighborhoods. *Schistosoma mansoni* was diagnosed using Kato-Katz (KK) and point-of-care circulating cathodic antigen (POC-CCA) tests. The Alcohol Use Disorders Identification Test (AUDIT) was used to determine the alcohol dependence score (ADS). Sociodemographic, behavioral, and environmental factors were also recorded. The prevalence of *S. mansoni* was 19.6% (95% CI: 15.7–24.0) by KK and 66.1% (95% CI: 58.8–73.3) by POC-CCA, yielding an overall prevalence of 34.6% (95% CI: 30.2 – 39.0). Diagnostic concordance between KK and POC-CCA was moderate (κ = 0.458; p < 0.001). Alcohol misuse prevalence was 54.3% (95% CI: 49.4–58.7) and schistosomiasis–alcoholism comorbidity prevalence was 17.4% (95% CI: 13.9 – 21.1). Independent predictors of infection included being 18–39 years old (AOR = 2.01; p = 0.001), residing in the Baloua neighborhood (AOR = 2.64; p < 0.001), and regularly performing domestic chores at transmission sites (AOR = 1.83; p = 0.006). Risk factors for alcoholism were male sex (AOR = 2.48; p = 0.002), regular consumption of adulterated whisky (AOR = 26.84; p = 0.005) and beer (AOR = 5.81; p < 0.001) and motivational factors, including conformity (AOR = 4.80; p < 0.001), coping (AOR = 3.06; p = 0.005), and social motives (AOR = 2.33; p = 0.002).

**Conclusion/significance:** This study confirms *S. mansoni* endemicity in Makenene and reveals a high prevalence of alcohol misuse, highlighting the comorbidity. These findings call for integrated interventions targeting both schistosomiasis transmission and harmful alcohol use.

**Author Summary:** This study presents, for the first time, documented data on the comorbidity between *Schistosoma mansoni* infection and alcoholism in a rural area of Cameroon. Through parasitological, sociodemographic, and behavioral approaches, it revealed the prevalence of schistosomiasis and alcohol abuse comorbidity in Makenene. The risk factors for *S. mansoni* infection included younger age (18–39 years), residence in the nearest riverine community, and frequent performance of domestic chores at transmission sites. In parallel, alcohol misuse was strongly associated with male sex and with the regular consumption of adulterated whisky and beer, two widely available and low-cost beverages. The primary motivations for alcohol abuse were the pursuit of social acceptance, the facilitation of social interactions, and the alleviation of psychological distress. The co-occurrence of these two major health problems highlights the urgent need for integrated strategies that address alcohol abuse within schistosomiasis control programs. These findings provide a basis for developing targeted interventions to simultaneously reduce the burden of schistosomiasis and alcohol abuse in semi-rural and rural settings.

## Introduction

Schistosomiasis is one of the most prevalent neglected tropical diseases, caused by parasitic trematodes of the genus *Schistosoma* [1]. Populations are exposed through agricultural, domestic, occupational, or recreational activities that involve contact with freshwater contaminated by cercariae. Cercariae are free-swimming larvae released by freshwater snails, which serve as intermediate hosts after becoming infected by water contaminated with human excreta containing parasite eggs [1,2]. In 2022, preventive chemotherapy targeting school-age children and high-risk adults was required for schistosomiasis in 50 countries for a total of 264.3 million people, including Cameroon [3]. Endemic areas are concentrated in tropical and subtropical regions, particularly among impoverished communities lacking access to safe water and adequate sanitation [1,3,4]. Globally, more than 700 million people are at risk of infection, and approximately 230 million are currently infected. The disease burden is especially high in sub-Saharan Africa, which accounts for 85% of cases and the majority of severe forms, with an estimated 200,000 deaths annually [5].

In Cameroon, the National Programme for the Control of Schistosomiasis and Intestinal Helminthiasis (PNLSHI) estimated that over 2 million people were infected in 2020 [6]. Schistosomiasis occurs in all ten regions of the country, with three human-infecting species identified: *Schistosoma haematobium, Schistosoma mansoni,* and *Schistosoma guineensis*. The Centre region reports the highest prevalence of *S. mansoni* infection (9.49%), with certain hotspots showing much higher rates, such as Makenene, where prevalence was 41% in 2010 [7]. Control strategies of schistosomiasis rely primarily on large-scale praziquantel administration to school-age children, supported by health education, vector control, improved sanitation, safe water supply, and environmental management of transmission sites, all of which are critical to achieve sustainable interruption of transmission [1–4].

*Schistosoma mansoni* infection is exacerbated by associated comorbidities. Among these, alcohol abuse is of particular concern due to its potential synergistic impact on hepatic pathology, since the liver is the main target organ in both conditions [8–9]. Classified among the most widely used psychoactive substances worldwide, alcohol is consumed by 43% of the global population, with alcohol use disorders affecting more than 600 million people [10,11]. In 2019, alcohol consumption accounted for 2.6 million deaths, representing 4.7% of global mortality [11]. Cameroon ranked fourth among 49 African countries assessed in 2019 for per capita alcohol consumption. The average of 10.1 liters of pure alcohol per person aged 15 years and older was more than double the African average of 4.5 L/person/year [11].

Alcohol misuse is the leading cause of liver disease, driving progressive damage including steatosis, alcoholic hepatitis, fibrosis, cirrhosis, and hepatocellular carcinoma [9–12]. Like chronic alcoholism, *S. mansoni* infection severely compromises liver function. At the chronic stage, granulomatous inflammation induced by parasite eggs leads to periportal fibrosis, often associated with splenomegaly and portal hypertension [8,13]. Concomitant alcohol intoxication may exacerbate schistosomiasis-related hepatic damage, amplifying clinical and pathophysiological consequences at the systemic level.

Therefore, sustainable control of schistosomiasis requires an approach that integrates alcohol consumption patterns in endemic populations with the identification of risk behaviors underlying persistent transmission. Based on this rationale, the present study aimed to determine the prevalence of schistosomiasis–alcoholism comorbidity among adults in an endemic focus of *S. mansoni* in Cameroon, as well as to identify the main risk factors associated with both conditions.

## Materials and methods

### Ethical statement

The study was approved by the Regional Committee of Research Ethics for Human Health of the Centre region, under the authority of the Ministry of Public Health of Cameroon, with ethical clearance issued under reference number CE N° 213/CRERSHC/2022. Authorization to conduct the study was also granted by the Ndikiniméki Health District, which oversees the Makenéné health area (N° 005-21/AE/MSP/DRSPC/DSN). The field survey was carried out in the communities with the agreement of local administrative and traditional authorities. After a detailed explanation of the study objectives, procedures, and the potential risks and benefits related to research and participants’ health, each adult residing in the target communities was free to decide whether or not to participate. Each participant signed an informed consent form. Data were anonymized during analysis to ensure participants’ confidentiality. The results of parasitological and immunological tests were individually delivered, and all *S. mansoni*-positive participants received, free of charge, a single dose of praziquantel (40 mg/kg body weight), in accordance with WHO recommendations [1]. Finally, all participants were sensitized to risk behaviors associated with schistosomiasis transmission and to the harmful effects of alcohol abuse.

### Study area

The study was conducted in Makenene, a semi-rural city in the Mbam and Inoubou Division, Centre Region of Cameroon. Located about 200 km northwest of Yaounde, Makenene covers an area of 885 km² and has an estimated population of 16,000 inhabitants. Schistosomiasis transmission in this area is favored by a large hydrographic network that includes several rivers, notably the Mock, Makombe, Makongo, and Makenene rivers, as well as numerous streams that mainly flow into the Mock River and the Noun River, which borders the city. It also encompasses swampy areas, particularly in the Baloua neighborhood, characterized by persistent humidity throughout the year due to the proximity of the water table. The local population is primarily engaged in market gardening and off-season crop farming.

The Mock River is the principal site of schistosomiasis transmission, hosting a high density of intermediate host snails of *S. mansoni* (*Biomphalaria pfeifferi*), thereby sustaining the parasite’s life cycle [14, 15]. *S. mansoni* infection has been reported in Makenene for more than three decades, with prevalence rates ranging from 82% in 1987 [16] to 41% in 2010 [7]. Among the sixteen villages constituting the geographical area of Makenene, three located along the Mock River were selected for this survey: Mock Centre, Mock Sud, and Carrière. The Baloua neighborhood, situated within the village of Mock Centre, was investigated separately. Baloua is located too close to the Mock River, and its high endemicity has been reported in several previous studies [14, 15, 17].

### Study design and population

A community-based and cross-sectional study was conducted in May 2022. In the sampled sites, *S. mansoni* infection affected individuals of both sexes and all age groups. Since the legal drinking age in Cameroon is 18 years, the study was restricted to individuals aged 18 years and above. The minimum sample size was calculated using the following formula: N= t^2^ *p *(1-p) / m where “N” represents the sample size, “t” the coefficient corresponding to a 95% confidence interval (1.96), “p” the prevalence of intestinal schistosomiasis in Makenene estimated at 41% at 2010 [7], and “m” the margin of error set at 5%. This approach resulted in a minimum sample size of 372 participants, corresponding to an expected number of at least 93 individuals per community site.

### Questionnaire

A structured digital interview questionnaire was used to collect data on sociodemographic characteristics, alcohol consumption level, and behaviors associated with both schistosomiasis risk and alcohol abuse. The survey was uploaded to the Survey 123 platform and administered by field epidemiologists who were familiar with both the questionnaire and digital data collection procedures. Responses were monitored in real time, with field supervisors ensuring data completeness and accuracy.

### Socio-environmental characteristics

The socio-environmental variables analyzed included age, sex, educational level, primary occupation, community of residence, and the relative distance between the place of residence and the transmission site, given their potential role in exposure to both parasitic and behavioral risk factors.

### Assessment of alcohol consumption level

Alcohol consumption was evaluated using the Alcohol Use Disorders Identification Test (AUDIT), developed by the World Health Organization [18]. The responses to the AUDIT items are rated on a 4-point Likert scale ranging from 0 to 4, yielding a maximum possible score of 40. The quantitative score provided by this standardized instrument (AUDIT) reflects the level of alcohol consumption and allows the classification of participants into four categories: non-use (AUDIT score = 0), low-risk use (AUDIT score = 1–6), harmful use (AUDIT score = 7–12), and alcohol dependence (AUDIT score ≥ 13) [19]. Based on the potentially injurious effects of alcohol on health, a binary categorization was applied, distinguishing abusive consumption (AUDIT score ≥ 7) from non-abusive consumption (AUDIT score ≤ 6).

### Determination of risk behaviors for *Schistosoma mansoni* infection

Several lifestyle habits likely to facilitate the transmission of schistosomiasis were recorded, including performing household chores and fishing at the transmission site, as well as, their respective frequencies. Participants were also asked about their knowledge of the disease, previous treatment with praziquantel, and the occurrence of symptoms suggestive of infestation during contact with transmission sites (e.g., itching or skin rash upon water exposure).

### Determination of factors associated with alcohol abuse

Drinking habits and motivations related to alcohol consumption were examined as potential determinants of abuse. The analysis considered both the type of beverage consumed and the frequency of intake. Alcoholic beverages identified in this study included beer, wine (red or white), spirits, and adulterated whisky, which are industrial products. Among traditional beverages, palm wine and two local drinks, “cha” (a maize turbid beer) and “odontol” (made from palm wine, sugar, and a tree bark) were considered.

Alcohol consumption motives were assessed using a series of eight specific questions posed to participants. Each item aimed to identify a particular reason for drinking alcohol (e.g., seeking pleasure, coping with stress, or socializing). To facilitate structured analysis within a validated conceptual framework, these motives were then grouped according to Cooper’s four-factor model [20]. This model classifies motives into social motives (drinking to enhance or facilitate social interactions), coping motives (drinking to reduce negative emotions or deal with psychological difficulties), enhancement motives (drinking to seek positive sensations and increase pleasure), and conformity motives (drinking driven by social pressure or the need for acceptance in a group).

### Diagnosis of *Schistosoma mansoni*

#### Sample collection

Following questionnaire administration, stool and urine samples were collected from each participant at their household. Participants were informed about the collection procedures and provided with two plastic containers to separately collect stool and urine. A unique identification code was affixed to each container and the corresponding survey form to ensure sample traceability. Urine samples were analyzed directly on site, while stool samples were transported to the laboratory of the Makenene Sub-district Health Center for *S. mansoni* diagnosis. All analyses were performed within a maximum of 24 hours after collection.

From a total of 431 participants, 383 provided stool samples for *S. mansoni* diagnosis using the Kato-Katz technique, and 165 provided urine samples for the POC-CCA test. Among them, 117 participants were simultaneously assessed with both diagnostic methods (Fig 1). A participant was considered infected with *S. mansoni* when she/he was diagnosed positive by at least one of the diagnostic methods.

**Fig 1.**
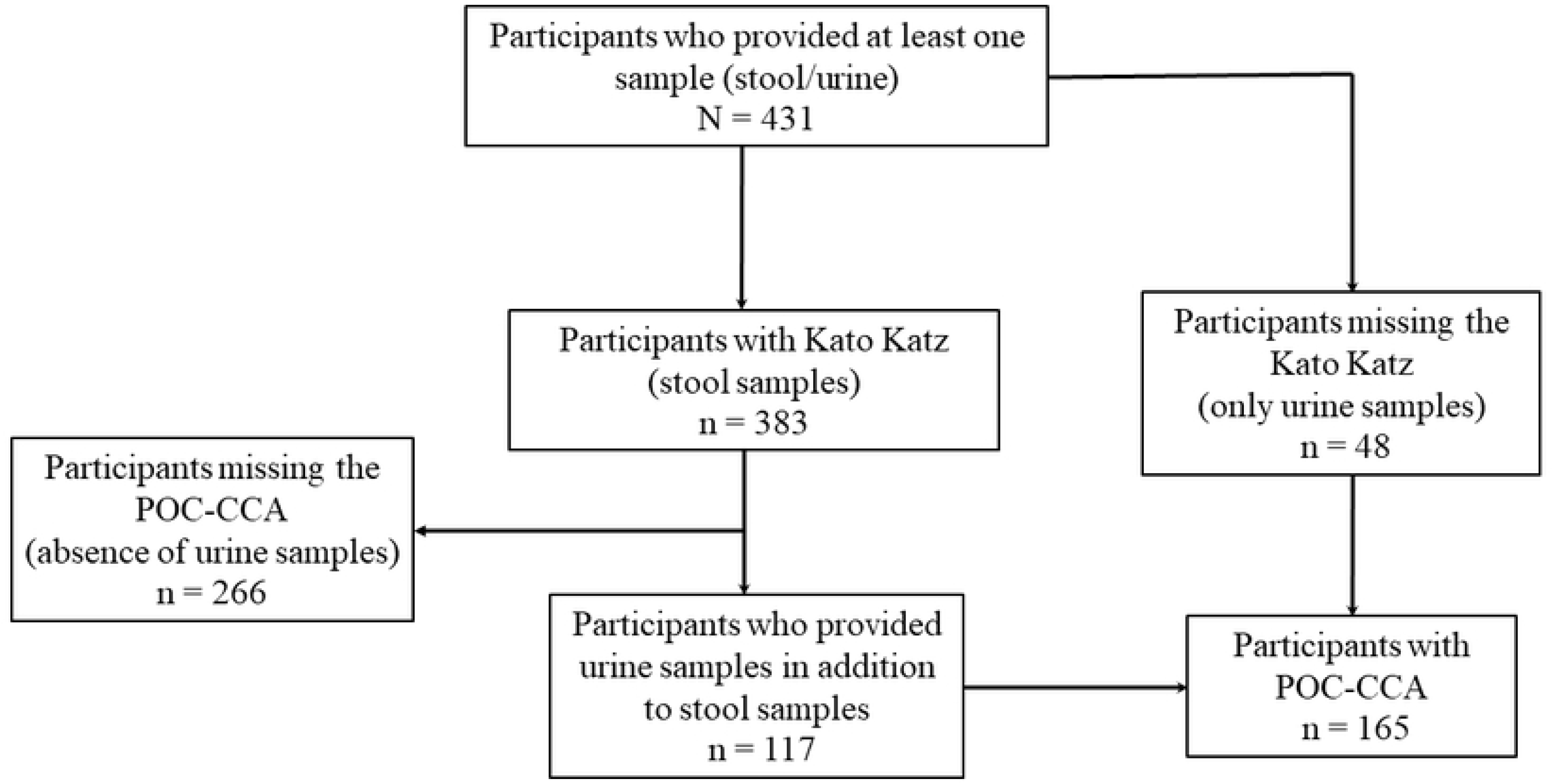
Participants flowchart for the diagnosis of *Schistosoma mansoni* infection.

### Detection of *Schistosoma mansoni* infection using the Kato-Katz method

Each stool sample was examined using a single thick smear prepared according to the Kato-Katz technique, with a standardized template of 41.7 mg [21]. After homogenization, a portion of the stool was sieved through a 212 µm mesh screen using a spatula. The sieved material was then transferred into a single-use template, mounted on a glass slide, and covered with a rectangle of cellophane soaked in Kato solution. The smear was obtained by inverting the slide onto a smooth surface and applying gentle pressure to evenly spread the sample.

The slides were examined under a light microscope within 12 h of preparation. A participant was considered positive when *S. mansoni* eggs, identifiable by their characteristic oval shape and lateral spine, were detected. Infection intensity was quantified by multiplying the total number of eggs observed per slide by a factor of 24 and expressed as a geometric mean of eggs per gram of stool (EPG). Positive samples were categorized into three groups according to infection level: light (1 – 99 EPG), moderate (100–399 EPG), and heavy (≥ 400 EPG) [1].

### Detection of circulating cathodic antigens of *Schistosoma mansoni*

The detection of circulating cathodic antigens (CCA) of *S. mansoni* was performed on freshly collected urine samples (within 15 min post-collection) using a point-of-care rapid diagnostic test (POC-CCA; Rapid Medical Diagnostics, Pretoria, South Africa, lot n° 211110105), administered directly at the participants’ households.

According to the manufacturer’s instructions, two drops of the urine sample were put into the circular well of the POC-CCA cassette. After complete absorption of the sample for exactly 20 min, the cassette was visually inspected. A test was considered valid if the control line turned a dark pink color. Otherwise, the sample was retested using a new cassette. A test was considered positive when, in addition to the control line, a test line was also visible.

### Statistical analysis

The collected data were exported in Excel format and then transferred to SPSS software version 27 for analysis. Qualitative variables were expressed as percentages, with confidence intervals (CI) provided for prevalence estimates, while quantitative data were expressed as means with their corresponding standard deviations. The Chi-square test (χ²) was used to compare prevalence between groups. Participants were classified into three main age groups: young adulthood (18 – 39 years old), middle adulthood (50 – 59 years old) and old age (≥ 60 years old). Student’s t-tests for non-independent data and one-way analysis of variance (ANOVA) were applied to compare the means of two groups and more than two groups, respectively.

The sensitivity, specificity, and accuracy of each diagnostic test were evaluated following the method described by Baratloo *et al*. [22]. The kappa coefficient (κ) and its 95% confidence interval (CI) were used to assess the concordance between Kato-Katz and POC-CCA, and the interpretation of κ values was performed according to the scale values: ≤ 0 as no agreement, 0.01–0.20 as slight, 0.21–0.40 as fair, 0.41– 0.60 as moderate, 0.61–0.80 as substantial and 0.81–1.00 as almost perfect agreement [23].

A binary logistic regression analysis was also performed using a stepwise backward elimination method to assess associations between variables. Odds ratios were estimated along with their 95% confidence intervals. Initially, a univariate analysis was conducted to examine the association between each independent variable and the outcomes of interest. Variables with a *p*-value < 0.05 and a defined OR at this stage were included in the binary logistic regression model to identify factors independently associated with *S. mansoni* infection as well as alcohol misuse. For all analyses, the level of statistical significance was set at 0.05.

## Results

### Socio-environmental characteristics of the study population

A total of 431 participants were included in the study, consisting of 199 women (46.2%) and 232 men (53.8%), yielding a sex ratio of 1.17. The population was predominantly composed of farmers and livestock breeders (61.7%), with the majority residing within 500 m of the main schistosomiasis transmission site (70.5%). Regarding educational attainment, most participants had reached, but not necessarily completed, secondary-level education, reflecting a generally low-to-moderate level of schooling (54.3%). The age distribution showed that nearly half of the participants (45.5%) were young adults aged 18–39 years. The overall mean age of the study population was 42.6 ± 15.7 years (95% CI: 41.1–44.1), ranging from 18 to 90 years (Table 1).

**Table 1.** Socio-environmental description of the population.

| **Variables** | **Modalities** | **Sample size**<br>**(N = 431)** | **Frequency (%)** |
| --- | --- | --- | --- |
| **Community** | Baloua | 115 | 26.7 |
|  | Carrière | 131 | 30.4 |
|  | Mock Centre | 103 | 23.9 |
|  | Mock Sud | 82 | 19.0 |
| **Sex** | Male | 232 | 53.8 |
|  | Female | 199 | 46.2 |
| **Age groups (years)** | 18 – 39 | 196 | 45.5 |
|  | 40 – 59 | 162 | 37.6 |
|  | ≥ 60 | 73 | 16.9 |
| **Level of education** | None | 35 | 8.1 |
|  | Primary | 146 | 33.9 |
|  | Secondary | 234 | 54.3 |
|  | Higher | 16 | 3.7 |
| **Main occupation** | Farmer/Breeder | 266 | 61.7 |
|  | Housewife | 61 | 14.2 |
|  | Technician | 33 | 7.7 |
|  | Student | 22 | 5.1 |
|  | Salesperson | 21 | 4.9 |
|  | Unemployed/Retired | 10 | 2.3 |
|  | Other | 18 | 4.2 |
| **Distance from**<br>**transmission site**<br>**(meters)** | < 500 | 304 | 70.5 |
|  | 500 – 1000 | 76 | 17.6 |
|  | > 1000 | 51 | 11.8 |

### Prevalence and severity of *Schistosoma mansoni* infection

#### Prevalence and severity of infection in the overall study population

The POC-CCA method revealed that 109 of 165 participants were positive for *S. mansoni* infection, giving a prevalence of 66.1% (95% CI: 58.8–73.3). The Kato-Katz method showed a prevalence of 19.6% (95% CI: 15.7–24.0), corresponding to 75 *S. mansoni*-positive participants over 383. Based on the fact that any participant diagnosed positive by at least one of the two diagnostic methods was considered infected with *S. mansoni*, out of the 431 participants examined, 149 were found to be infected, corresponding to an overall prevalence of 34.6% (95% CI: 29.7– 39.0) (Table 2).

**Table 2.** Overall prevalence of *Schistosoma mansoni* infection.

|  | POC-CCA | Kato Katz | Overall prevalence |
| --- | --- | --- | --- |
| **Number examined** | 165 | 383 | 431 |
| **Number of positive** | 109 | 75 | 149 |
| **Prevalence (%)** | 66.1 | 19.6 | 34.6 |
| **95% CI** | 58.8 – 73.3 | 15.7 – 24.0 | 29.9 – 39.0 |
| ***p*** | < 0.001 |  |  |

95% CI: 95% confidence interval associated with prevalence.

The severity of infection, assessed based on the mean egg load among participants diagnosed positive by the Kato-Katz method (n = 75), was 100.3 ± 2.9 EPG (95% CI: 78.5 – 128.3), corresponding to a moderate infection intensity in the overall infected population; however, values ranged from 24 to 4,440 EPG. According to the WHO stratification, infection was classified as light in 57.3% of cases, moderate in 33.3%, and heavy in 9.3% (Table 3).

**Table 3.** Infection intensity of *Schistosoma mansoni*-positive participants.

|  | Low intensity | Moderate intensity | Heavy intensity | Overall intensity |
| --- | --- | --- | --- | --- |
| **n<sub>kk</sub> (%)** | 43 (57.3%) | 25 (33.3%) | 7 (9.3) | 75 |
| **Mean EPG ± SD** | 48.3 ± 1.6 | 189.6 ± 1.4 | 918.5 ± 2.2 | 100.3 ± 2.9 |
| **95% CI** | 41.5 – 56.4 | 163.5 – 219.8 | 446.1 – 1891.2 | 78.5 – 128.3 |
| ***p*** | < 0.001 |  |  |  |

n<sub>kk</sub>: Schistosomiasis cases diagnosed by Kato-Katz; EPG: Egg per gram of stool; SD: Standard deviation; 95% CI: 95% confidence interval associated with the mean of infection intensity; Low intensity = <100 EPG; Moderate intensity = 100–400 EPG; Heavy intensity = >400 EPG.

#### Diagnostic performance of the Kato-Katz method compared to the POC-CCA method

Out of a total of 117 participants diagnosed with both diagnostic methods, concordant results between KK and POC-CCA were observed in 83 (70.9%) people, among whom 35 (29.9%) were positive and 48 (41.0%) negative with both tests. Discordant results were recorded in 34 (29.1%) participants, all of whom tested positive with POC-CCA but negative with Kato-Katz (Table 4).

**Table 4.** Agreement between Kato-Katz and POC-CCA methods in the diagnosis of *Schistosoma mansoni* infection.

| | | POC-CCA | | Total | $\kappa$ ; $p$ (kappa) |
| --- | --- | --- | --- | --- | --- |
|  |  | Positive | Negative |  |  |
| Kato-Katz | Positive | 35 | 0 | 35 | 0.458 ; < 0.001 |
|  | Negative | 34 | 48 | 82 |  |
|  | Total | 69 | 48 | 117 |  |

Accuracy: $((35 + 48) / 117) \times 100 = 70.9\%$
Sensitivity (POC-CCA): $(35 / 35) \times 100 = 100\%$
Specificity (POC-CCA): $(48 / (34 + 48)) \times 100 = 58.5\%$
Sensitivity (Kato-Katz) = $(35/69) \times 100 = 50.7\%$
Specificity (Kato-Katz) = $(48/48) \times 100 = 100\%$

Performance indicators calculated from the contingency matrix showed that the sensitivity of the POC-CCA test was 100%, reflecting its ability to detect all positive cases. Its specificity was 58.5%, indicating a relatively high number of false-positive results.

For the Kato-Katz test, the sensitivity was 50.7%, showing a moderate ability to detect positive cases, while the specificity was 100%, meaning no uninfected person was wrongly identified as positive.

The diagnostic concordance index, expressed by the Kappa coefficient, was 0.458 (95% CI: 0.33–0.59) with *p* < 0.001, indicating a moderate agreement between the two diagnostic methods, but statistically significant.

Overall, these findings highlight the significantly higher diagnostic performance of the POC-CCA method relative to the Kato-Katz method.

#### Prevalence and severity of *Schistosoma mansoni* infection according to socio-demographic characteristics

To assess the distribution of the prevalence and severity of *S. mansoni* infection, data were stratified by community, sex, and age intervals (Table 5). At the community level, Baloua had the highest prevalence of 50.4% (95% CI: 41.0–59.3), followed by Carrière with a 33.6% (95% CI: 25.7–42.4) prevalence. Lower prevalence rates were observed in Mock Sud (28.0%; 95% CI: 18.7– 38.7) and Mock Centre (23.3%; 95% CI: 15.5–31.3). A significant difference was observed between those two communities and Baloua (*p* < 0.001). The parasite burden, which reflects infection severity, was 88.4 ± 3.2 EPG and 90.9 ± 2.5 EPG in Carrière and Mock Centre, respectively, reflecting a light infection intensity. A moderate infection intensity was recorded in Baloua (109.4 ± 3.1 EPG) and Mock Sud (146.4 ± 2.2 EPG). However, no significant difference was observed at the community level (*p* = 0.795).

**Table 5.** Distribution and severity of *Schistosoma mansoni* infection according to sociodemographic criteria Variable.

| Variable | Prevalence of *S. mansoni* |  |  |  | Intensity of infection |  |  |  |
| --- | --- | --- | --- | --- | --- | --- | --- | --- |
|  | N = 431 | n (%) | 95% CI | *p* | n<sub>kk</sub> = 75 | Mean EPG ± SD | 95% CI | *p* |
| Community |  |  |  |  |  |  |  |  |
| Baloua | 115 | 58 (50.4)<sup>ms, mc</sup> | 41.0 - 59.3 | < 0.001 | 33 | 109.4 ± 3.1 | 73.0 – 163.7 | 0.795 |
| Carrière | 131 | 44 (33.6) | 25.7 - 42.4 |  | 21 | 88.4 ± 3.2 | 52.0 – 150.2 |  |
| Mock Centre | 103 | 24 (23.3)<sup>b</sup> | 15.5 - 31.3 |  | 17 | 90.9 ± 2.5 | 57.3 – 144.2 |  |
| Mock Sud | 82 | 23 (28.0)<sup>b</sup> | 18.7 - 38.7 |  | 4 | 146.4 ± 2.2 | 43.3 – 495.6 |  |
| Sex |  |  |  |  |  |  |  |  |
| Male | 232 | 72 (31.0) | 25.0 – 37.1 | 0.096 | 36 | 71.7 ± 2.8 | 50.9 – 101.1 | 0.008 |
| Female | 199 | 77 (38.7) | 31.7 – 46.2 |  | 39 | 136.8 ± 2.8 | 97.8 – 191.3 |  |
| Age groups (years) |  |  |  |  |  |  |  |  |
| 18 – 39 | 196 | 87 (44.4)<sup>β, γ</sup> | 37.8 – 51.4 | < 0.001 | 36 | 113.8 ± 3.4 | 75.5 – 171.4 | 0.032 |
| 40 – 59 | 162 | 46 (28.4)<sup>α</sup> | 21.0 – 35.4 |  | 28 | 92.1 ± 2.6 | 63.9 – 132.6 |  |
| ≥ 60 | 73 | 16 (21.9)<sup>α</sup> | 12.9 – 31.4 |  | 11 | 82.8 ± 2.5 | 45.0 – 152.3 |  |

N: Total sample size; n: Schistosomiasis cases; n□□: Schistosomiasis cases diagnosed by Kato-Katz; EPG: Egg per gram of stool; SD: Standard deviation; 95% CI: 95% confidence interval associated with the prevalence or the mean of infection intensity.
b: significantly different from Baloua; c: significantly different from Carrière; mc: significantly different from Mock Centre; ms: significantly different from Mock Sud. α: significantly different from 18–39 years; β: significantly different from 40–59 years; γ: significantly different from ≥ 60 years.

By sex, prevalence tended to be higher among women (38.7%) than men (31.0%), although this difference did not reach statistical significance (*p* = 0.096). Likewise, a significant difference in parasite burden was recorded between sex (*p* = 0.008). In men, the parasite burden fell into the light infection intensity category (71.7 ± 2.8 EPG), while for women, it corresponded to moderate infection intensity (136.8 ± 2.8 EPG) (Table 5).

Across age groups, prevalence decreased significantly with increasing age (*p* < 0.001). Participants aged 18–39 years had the highest prevalence of 44.4% (95% CI: 37.8–51.4), compared to 28.4% (95% CI: 21.0–35.4) for the 40–59 years and to 21.9% (95% CI: 12.9–31.4) for participants aged ≥ 60 years. Similarly, mean egg counts significantly decreased with age, from 113.8 ± 3.4 EPG for the 18–39 years participants to 82.8 ± 2.5 eggs/g among those aged ≥ 60 years (*p* = 0.032). Thus, only the 18-39 years’ age group had a moderate infection intensity, with the other age groups falling into the light intensity category (Table 5).

### Prevalence of alcohol consumption levels

#### Prevalence of alcohol consumption levels in the general population

The distribution of participants according to their level of alcohol consumption (Table 6) revealed a markedly unequal pattern within the study population (*p* < 0.001). The majority of the participants, 42.9% (95% CI: 38.3–47.3), were classified in the “low-risk use” category (AUDIT = 1–6). The “harmful use” category (AUDIT = 7–12) was represented by 28.8% (95% CI: 24.6– 32.9) of the participants, while 25.5% (95% CI: 21.6–29.7) of the participants fell in the “alcohol dependence” category (AUDIT ≥ 13). In contrast, the non-consumers of alcohol, the “non-use” category (AUDIT = 0), represented 2.8% (95% CI: 1.4–4.4) (Table 6). The mean AUDIT score among all alcohol consumers (AUDIT ≥ 1, n = 419) was 8.9 ± 6.9 (95% CI: 8.3–9.6), with scores ranging from 1 to 30. Consistently, it increased significantly across categories (*p* < 0.001): 2.9 ± 1.7 for low-risk use, 9.5 ± 1.7 for harmful use, and 18.5 ± 4.56 for alcohol dependence (Table 6).

**Table 6.** Levels of alcohol consumption in the general population.

| Variables | Prevalence<br>n (%) | 95% CI | *p* | AUDIT ± SD | 95% CI | *p* |
| --- | --- | --- | --- | --- | --- | --- |
| **Alcohol consumption level** |  |  |  |  |  |  |
| Non-use (AUDIT =0) | 12 (2.8) | 1.4 – 4.4 |  | 0 | / |  |
| Low-risk use (AUDIT =1 - 6) | 185 (42.9) | 38.3 – 47.3 | < | 2.9 ± 1.7 | 2.7 – 3.2 | < |
| Harmful use (AUDIT =7 - 12) | 124 (28.8) | 24.6 – 32.9 | 0.001 | 9.5 ± 1.7 | 9.1 – 9.8 | 0.001 |
| Alcohol dependence (AUDIT ≥ 13) | 110 (25.5) | 21.6 – 29.7 |  | 18.5 ± 4.6 | 17.7 – 19.4 |  |
| **Use of alcohol** |  |  |  |  |  |  |
| Non-abuse (AUDIT ≤ 6) | 197 (45.7) | 40.1 – 50.1 | 0.074 | 2.3 ± 1.7 | 2.1 – 2.5 | < |
| Abuse (AUDIT ≥ 7) | 234 (54.3) | 49.4 – 58.7 |  | 13.7 ± 5.6 | 13.0 – 14.4 | 0.001 |

AUDIT: Alcohol Use Disorder Identification Test score; it represents the mean score of the participants in each group. n (%): number of participants (n) and prevalence (%); 95% CI: 95% confidence interval associated with the prevalence or the AUDIT score; SD: standard deviation.

Overall, the prevalence of alcohol abuse (AUDIT ≥ 7) in the study population was 54.3% (95% CI: 49.4–58.7), whereas it was 45.7% (95% CI: 40.1–50.1) for the non-abuse of alcohol (AUDIT ≤ 6). However, a significant difference in terms of alcohol consumption intensity was recorded between these groups (*p* < 0.001), with mean AUDIT scores of 13.7 ± 5.63 (95% CI: 13.0–14.4) among alcohol abuse consumers compared to 2.32 ± 1.73 (95% CI: 2.1–2.5) among non-abuse alcohol consumers (Table 6).

#### Prevalence of alcohol abuse consumption according to socio-demographic criteria

To assess the distribution of alcohol abuse consumption across different sociodemographic variables, prevalence rates and mean consumption scores were analyzed within groups defined by community of origin, sex, and age (Table 7).

**Table 7.** Association between alcohol consumption levels and sociodemographic criteria.

| Variable | N = 431 | Prevalence of “alcohol abuse” n (%) | 95% CI | *p* | AUDIT ± SD | 95% CI | *p* |
| --- | --- | --- | --- | --- | --- | --- | --- |
| Community |  |  |  |  |  |  |  |
| Baloua | 115 | 47 (40.9) <sup>c, ms</sup> | 32.2 – 50.4 | < 0.001 | 6.6 ± 5.7 <sup>c, ms</sup> | 5.6 – 7.7 | < 0.001 |
| Carrière | 131 | 91 (69.5) <sup>b</sup> | 61.1 – 77.1 |  | 11.4 ± 7.8 <sup>b, mc</sup> | 10.0 – 12.7 |  |
| Mock Centre | 103 | 41 (39.8) <sup>c, ms</sup> | 31.1 – 49.5 |  | 6.6 ± 6.2 <sup>c, ms</sup> | 5.4 – 7.8 |  |
| Mock Sud | 82 | 55 (67.1) <sup>b, mc</sup> | 56.1 – 76.8 |  | 10.0 ± 6.3 <sup>b</sup> | 8.6 – 11.3 |  |
| Sex |  |  |  |  |  |  |  |
| Male | 232 | 161 (69.4) | 63.8 – 75.3 | < 0.001 | 11.3 ± 7.4 | 10.3 – 12.2 | < 0.001 |
| Female | 199 | 73 (36.7) | 30.0 – 43.3 |  | 5.7 ± 5.0 | 5.0 – 6.4 |  |
| Age groups (years) |  |  |  |  |  |  |  |
| 18 – 39 | 196 | 98 (50.0) <sup>β</sup> | 42.9 – 57.1 | 0.032 | 8.1 ± 7.0 | 7.1 – 9.1 | 0.115 |
| 40 – 59 | 162 | 101 (62.3) <sup>α, γ</sup> | 54.3 – 69.8 |  | 9.6 ± 7.0 | 8.5 – 10.7 |  |
| ≥ 60 | 73 | 35 (47.9) <sup>β</sup> | 37.0 – 60.3 |  | 8.2 ± 6.7 | 6.7 – 9.8 |  |

N: Total sample size; n (%): Cases of alcohol abuse (n) and prevalence (%); 95% CI: 95% confidence interval associated with prevalence or the AUDIT score; SD: Standard deviation; AUDIT: Alcohol Use Disorder Identification Test score; it represents the mean score of the participants in each group.
b: significantly different from Baloua; c: significantly different from Carrière; mc: significantly different from Mock Centre; ms: significantly different from Mock Sud. α: significantly different from 18–39 years; β: significantly different from 40–59 years; γ: significantly different from ≥ 60 years.

Community-level analysis revealed significant differences (*p* < 0.001) in both the prevalence and alcohol consumption intensity. The Carrière and Mock Sud communities displayed the highest prevalence rates of alcohol abuse, 69.5% (95% CI: 61.1–77.1) and 67.1% (95% CI: 56.1–76.8), respectively, while Baloua and Mock Centre exhibited much lower prevalences, around 40%. This disparity was also reflected in the mean alcohol consumption intensity (AUDIT score), which was higher in Carrière (11.4 ± 7.8) and Mock Sud (10.0 ± 6.3), corresponding to alcohol abuse consumption (AUDIT ≥ 7). In contrast, lower scores in Baloua (6.6 ± 5.7) and Mock Center (6.6 ± 6.2), consistent with non-abuse alcohol consumption (AUDIT ≤ 6).

Concerning sex, the prevalence of alcohol abuse consumption was higher in men (69.4%) than in women (36.7%) (*p* < 0.001), with an AUDIT score of 11.3 ± 7.4 among men and 5.7 ± 5.0 for women, categorizing men as alcohol abuse consumers and women as non-abuse alcohol consumers.

Across all age groups, the AUDIT score did not differ significantly (*p* = 0.115) and was confirmed as the “alcohol abuse” category. However, the prevalence of alcohol abuse consumption was higher among participants aged 40-59 years (62.3% (95% CI: 54.3–69.8)) than among others (*p* = 0.032).

### Prevalence of schistosomiasis–alcoholism comorbidity

Schistosomiasis–alcoholism comorbidity was defined as the concomitant presence of *S. mansoni* infection and alcohol abuse consumption (AUDIT ≥ 7) within the same individual. The mean intensity of alcohol consumption (AUDIT score) and the intensity of schistosomal infection (EPG) among co-morbid participants are provided.

#### Prevalence of the comorbidity in the general population

As shown in Table 8, the overall prevalence of schistosomiasis–alcohol comorbidity was 17.4% (95% CI: 13.9–21.1). The mean parasitic burden was 95.4 ± 3.0 (95% CI: 66.8–136.4) within co-morbid participants, corresponding to a light *S. mansoni* infection intensity. The AUDIT score among those participants was 13.0 ± 5.5 (95% CI: 11.8–14.3), reflecting an alcohol dependence.

**Table 8.** Prevalence of schistosomiasis *mansoni*-alcoholism comorbidity in the global population and according to sociodemographic criteria.

|  |  | Prevalence of comorbidity |  |  | Intensity of alcohol abuse |  |  | Severity of *S. mansoni* infection |  |  |  |
| --- | --- | --- | --- | --- | --- | --- | --- | --- | --- | --- | --- |
| Variable | N | Prevalence<br>n (%) | 95% CI | *p* | AUDIT ± SD | 95% CI | *p* | n<sub>kk</sub> | Mean EPG ± SD | 95% CI | *p* |
| Community |  |  |  |  |  |  |  |  |  |  |  |
| Baloua | 115 | 27 (23.5)<sup>mc</sup> | 16.5 – 31.3 | 0.001 | 12.6 ± 5.0 | 10.9 – 14.6 | 0.334 | 16 | 134.0 ± 3.4 | 69.4 – 258.9 | 0.037 |
| Carrière | 131 | 31 (23.7)<sup>mc</sup> | 16.0 – 31.3 |  | 14.0 ± 6.2 | 11.9 – 16.4 |  | 15 | 71.1 ± 3.0 | 38.9 – 129.8 |  |
| Mock Centre | 103 | 8 (7.8)<sup>b,c</sup> | 2.9 – 13.6 |  | 10.1 ± 2.9 | 8.4 – 12.1 |  | 6 | 64.0 ± 1.8 | 35.1 – 116.6 |  |
| Mock Sud | 82 | 9 (11.0) | 4.9 – 18.3 |  | 13.0 ± 5.5 | 10.0 – 16.9 |  | 2 | 190.5 ± 1.19 | 38.6 – 940.3 |  |
| Sex |  |  |  |  |  |  |  |  |  |  |  |
| Male | 232 | 47 (20.3) | 15.1 – 25.4 | 0.091 | 13.7 ± 5.9 | 12.2 – 15.4 | 0.121 | 21 | 72.6 ± 3.1 | 43.4 – 121.3 | 0.007 |
| Female | 199 | 28 (14.1) | 9.5 – 19.1 |  | 11.7 ± 4.5 | 10.1 – 13.5 |  | 18 | 131.3 ± 2.7 | 79.5 – 216.8 |  |
| Age groups (years) |  |  |  |  |  |  |  |  |  |  |  |
| 18 - 39 | 196 | 43 (21.9)<sup>γ</sup> | 16.5 – 27.8 | 0.026 | 14.1 ± 5.9 | 12.4 – 16.1 | 0.112 | 18 | 119.2 ± 3.5 | 63.9 – 222.2 | 0.076 |
| 40 - 59 | 162 | 26 (16.0) | 10.2 – 22.1 |  | 11.3 ± 3.9 | 9.8 – 13.0 |  | 16 | 83.4 ± 2.6 | 49.9 – 139.6 |  |
| ≥ 60 | 73 | 6 (8.2)<sup>α</sup> | 2.5 – 14.7 |  | 12.0 ± 7.0 | 7.5 – 18.2 |  | 5 | 65.9 ± 2.6 | 20.1 – 215.6 |  |
| Global population | 431 | 75 (17.4) | 13.9 – 21.1 | - | 13.0 ± 5.5 | 11.8 – 14.3 | - | 39 | 95.4 ± 3.0 | 66.8 – 136.4 |  |

N: total sample size; n (%): cases of schistosomiasis-alcoholism comorbidity (n) and prevalence (%); 95% CI: 95% confidence interval associated with the prevalence or the mean of AUDIT score or the mean of infection intensity; AUDIT: Alcohol Use Disorder Identification Test score; it represents the mean score of the participants in each group; SD: Standard deviation; n□□: schistosomiasis mansoni cases diagnosed by Kato-Katz; EPG: Egg per gram of stool.
b: significantly different from Baloua; c: significantly different from Carrière; mc: significantly different from Mock Centre.
α: significantly different from the 18–39 years; γ: significantly different from the ≥ 60 years.

#### Prevalence of the comorbidity according to sociodemographic criteria

At the community level, a significant variation of the comorbidity prevalence was recorded across communities (*p* = 0.001). Baloua and Carrière displayed higher prevalence of 23.5% (95% CI: 16.5–31.3) and 23.7% (95% CI: 16.0–31.3), respectively, whereas Mock Centre and Mock Sud exhibited lower prevalence of 7.8% (95% CI: 2.9–13.6) and 11.0% (95% CI: 4.9–18.3), respectively. Regarding the intensity of alcohol consumption, no significant difference was observed between communities (p = 0.334). However, the AUDIT score indicated harmful drinking for co-morbid participants from Baloua (12.6 ± 5.0) and Mock Centre (10.1 ± 2.9), whereas alcohol dependence was registered for co-morbid participants from Carrière (14.0 ± 6.2) and Mock Sud (13.0 ± 5.5). Significant variability of *S. mansoni* infection intensity was recorded across communities (*p* = 0.037), with a moderate intensity of infection at Mock Sud (190.5 ± 1.19 EPG) and Baloua (134.0 ± 3.4 EPG) and a light intensity of infection at Carrière (71.1 ± 3.0 EPG) and Mock Centre (64.0 ± 1.8 EPG) (Table 8).

#### Prevalence of the comorbidity according to sex

The prevalence of schistosomiasis mansoni-alcoholism comorbidity was slightly higher among men, 20.3% (95% CI: 15.1–25.4), compared to 14.1% (95% CI: 9.5–19.1) among women, without a statistically significant variation (*p* = 0.091). Although the intensity of alcohol consumption did not differ significantly between sexes (*p* = 0.121), men had an AUDIT score of 13.7 ± 5.9, consistent with alcohol dependence, compared to 11.7 ± 4.5 for women, indicating harmful drinking. Regarding the severity of *S. mansoni* infection, women displayed moderate intensity of infection (131.3 ± 2.7 EPG) compared to light intensity of infection for men (72.6 ± 3.1 EPG) (*p* = 0.007) (Table 8).

#### Prevalence of the comorbidity according to age groups

The prevalence of schistosomiasis–alcohol comorbidity significantly decreased with age (*p* = 0.026). Participants aged 18–39 years showed the highest prevalence at 21.9% (95% CI: 16.5–27.8), followed by those aged 40–59 years at 16.0% (95% CI: 10.2–22.1), while the ≥60 years group had the lowest prevalence at 8.2% (95% CI: 2.5–14.7). The level of alcohol consumption was generally similar across age groups (*p* = 0.112), although slightly higher among young adults (14.1 ± 5.9), indicative of alcohol dependence, compared to middle-aged adults (11.3 ± 3.9) and older adults (12.0 ± 7.0), both suggesting harmful drinking behaviors. Regarding infection severity, a moderate intensity of infection (119.2 ± 3.5 EPG) was observed in the 18–39 years age group, while a light intensity of infection (65.9 ± 2.6 EPG) was recorded in older age groups (Table 8).

### Factors associated with *Schistosoma mansoni* infection

#### Sociodemographic factors of *Schistosoma mansoni* infection

Among the sociodemographic factors analyzed, young adults (18–39 years) were 2.8 times more at risk of *S. mansoni* infection (χ² = 10.843; *p* = 0.001). Similarly, residents of the Baloua neighborhood had a 2.5-fold higher risk (χ² = 17.452; *p* < 0.001), while living within 500 meters of a transmission site was associated with a twofold increase in risk (χ² = 4.186; *p* = 0.041). In contrast, living in Mock Centre was significantly associated with a lower risk of infection (χ² = 7.600; *p* = 0.006) (Table 9).

**Table 9.** Association between sociodemographic factors and *Schistosoma mansoni* infection.

| Variables | *S. mansoni*<br>positive | *S. mansoni*<br>negative | OR (95% CI) | $\chi^2$ ; *p* |
| --- | --- | --- | --- | --- |
|  | N=149 ; n (%) | N=282 ; n (%) |  |  |
| **Sex** |  |  |  |  |
| Male | 72 (31.0) | 160 (69.0) | 0.71 (0.47-1.06) | 2.778 ; 0.096 |
| Female | 77 (38.7) | 122 (61.3) | 1 |  |
| **Age groups (years)** |  |  |  |  |
| 18-39 | 87 (44.4) | 109 (55.6) | 2.84 (1.52-5.29) | 10.843 ; 0.001 |
| 40-59 | 46 (28.4) | 116 (71.6) | 1.41 (0.73-2.70) | 1.081 ; 0.298 |
| ≥ 60 | 16 (21.9) | 57 (78.1) | 1 |  |
| **Level of education** |  |  |  |  |
| None/primary | 61 (33.7) | 120 (66.3) | 0.93 (0.62-1.40) | 0.104 ; 0.747 |
| Secondary/higher | 88 (35.2) | 162 (64.8) | 1 |  |
| **Main occupation** |  |  |  |  |
| Farmer/Breeder | 98 (36.8) | 168 (63.2) | 1.30 (0.86-1.97) | 1.585 ; 0.208 |
| Salesperson | 7 (33.3) | 14 (66.7) | 0.94 (0.37-2.39) | 0.015 ; 0.903 |
| Housewife | 19 (31.1) | 42 (68.9) | 0.83 (0.46-1.49) | 0.368 ; 0.544 |
| Student | 7 (31.8) | 15 (68.2) | 0.87 (0.35-2.20) | 0.078 ; 0.780 |
| Technician | 10 (30.3) | 23 (69.7) | 0.81 (0.37-1.75) | 0.288 ; 0.592 |
| Unemployed/Retired | 2 (20.0) | 8 (80.0) | 0.46 (0.09-2.22) | 0.961 ; 0.327 |
| **Community** |  |  |  |  |
| Baloua | 58 (50.4) | 57 (49.6) | 2.51 (1.62-3.90) | 17.452 ; < 0.001 |
| Carrière | 44 (33.6) | 87 (66.4) | 0.93 (0.60-1.44) | 0.080 ; 0.777 |
| Mock-Centre | 24 (23.3) | 79 (76.7) | 0.49 (0.29-0.82) | 7.600 ; 0.006 |
| Mock-Sud | 23 (28.0) | 59 (72.0) | 0.69 (0.40-1.17) | 1.904 ; 0.168 |
| **Household–transmission site distance (m)** |  |  |  |  |
| < 500 | 111 (36.5) | 193 (63.5) | 2.09 (1.03-4.24) | 4.186 ; 0.041 |
| 500 – 1000 | 27 (35.5) | 49 (64.5) | 2.00 (0.88-5.53) | 2.785 ; 0.095 |
| > 1000 | 11 21.6) | 40 (78.4) | 1 |  |

#### Behavioral factors associated with *Schistosoma mansoni* infection

Among the lifestyle-related factors examined, the regular performance of household chores at the transmission site was the only variable significantly associated with *S. mansoni* infection, doubling the risk of infection (χ² = 11.689; *p* = 0.001) (Table 10).

**Table 10.** Association between behavioral characteristics and *Schistosoma mansoni* infection.

| Variables | *S. mansoni*<br>positive<br>N=149 ; n (%) | *S. mansoni*<br>negative<br>N=282 ; n (%) | OR (95% CI) | $\chi^2$ ; $p$ |
| --- | --- | --- | --- | --- |
| **Alcohol abuse** |  |  |  |  |
| Yes | 75 (32.1) | 159 (67.9) | 0.78 (0.52-1.16) | 1.437 ; 0.231 |
| No | 74 (37.6) | 123 (62.4) | 1 |  |
| **Knowledge of schistosomiasis** |  |  |  |  |
| Yes | 133 (35.4) | 243 (64.6) | 1.33 (0.71-2.47) | 0.837 ; 0.360 |
| No | 16 (29.1) | 39 (70.9) | 1 |  |
| **Previous treatment with PZQ** |  |  |  |  |
| Never | 92 (37.4) | 154 (62.6) | 1 |  |
| More than 12 months | 42 (32.8) | 86 (67.2) | 0.81 (0.52-1.28) | 0.769 ; 0.381 |
| Less than 12 months | 15 (26.3) | 42 (73.7) | 0.59 (0.31-1.13) | 2.455 ; 0.117 |
| **Housework practices at the transmission site** |  |  |  |  |
| Never | 63 (28.0) | 162 (72.0) | 1 |  |
| More than 12 months | 12 (29.3) | 29 (70.7) | 1.06 (0.51-2.21) | 0.027 ; 0.868 |
| Less than 12 months | 74 (44.8) | 91 (55.2) | 2.09 (1.37-3.19) | 11.689 ; 0.001 |
| **Regular fishing practice** |  |  |  |  |
| Yes | 11 (42.3) | 15 (57.7) | 1.41 (0.63-3.17) | 0.732 ; 0.392 |
| No | 138 (34.1) | 267 (65.9) | 1 |  |
| **Itching (water contact)** |  |  |  |  |
| Never | 88 (34.2) | 169 (65.8) | 1 |  |
| Irregularly | 46 (33.8) | 90 (66.2) | 0.98 (0.63-1.52) | 0.006 ; 0.934 |
| Regularly | 15 (39.5) | 23 (60.5) | 1.25 (0.62-2.52) | 0.398 ; 0.528 |
| **Skin rash (water contact)** |  |  |  |  |
| Never | 117 (36.2) | 206 (63.8) | 1 |  |
| Irregularly | 27 (27.0) | 73 (73.0) | 0.65 (0.39-1.07) | 2.869 ; 0.090 |
| Regularly | 5 (62.5) | 3 (37.5) | 2.93 (0.68-12.50) | 2.119 ; 0.145 |

#### Multivariate analysis of *Schistosoma mansoni* infection and its associated factors

The binary logistic regression model revealed three factors independently associated with a higher risk of *S. mansoni* infection. Young adults (18–39 years) had a twofold higher risk of infection (AOR = 2.01; *p* = 0.001), while residing in the Baloua neighborhood was linked to a 2.6-fold increase in risk (AOR = 2.64; *p* < 0.001). Additionally, regular performance of household chores at the transmission site increased the likelihood of infection by 1.8 times (AOR = 1.83; *p* = 0.006) (Table 11).

**Table 11.** Factors independently associated with *Schistosoma mansoni* infection.

| Variables | Crude odd ratio |  |  | Adjusted odd ratio |  |
| --- | --- | --- | --- | --- | --- |
|  | *S. mansoni*<br>positive, n (%) | COR (95% CI) | *p* | AOR (95% CI) | *p* |
| **Age groups (years)** |  |  |  |  |  |
| 18-39 ans | 87 (44.4) | 2.03 (1.32-3.11) | 0.001 | 2.01 (1.31-3.08) | 0.001 |
| **Community** |  |  |  |  |  |
| Baloua | 58 (50.4) | 2.49 (1.53-4.08) | < 0.001 | 2.64 (1.67-4.16) | < 0.001 |
| Mock-Centre | 24 (23.3) | 0.77 (0.44-1.35) | 0.372 |  |  |
| **Household–transmission site distance (m)** |  |  |  |  |  |
| < 500 m | 111 (36.5) | 1.26 (0.77-2.05) | 0.352 | / | / |
| **Housework practices at the transmission site** |  |  |  |  |  |
| Regularly | 74 (44.8) | 1.70 (1.09-2.65) | 0.019 | 1.83 (1.19-2.82) | 0.006 |

### Factors associated with alcohol abuse

#### Sociodemographic factors associated with alcohol abuse

Among the sociodemographic factors analyzed, men were found to be at significantly higher risk of alcohol abuse, with a risk 3.9 times greater than that of women (χ² = 46.193; *p* < 0.001). Concerning age groups, only middle adults aged 40–59 years showed a significantly increased risk, being approximately 1.7 times more likely to alcohol abuse compared to other age categories (χ² = 6.274; *p* = 0.009) (Table 12).

**Table 12.** Association between sociodemographic factors and alcohol abuse.

| Variables | Alcohol abuse<br>N=234 ; n (%) | Non-abuse of<br>alcohol<br>N=197 ; n (%) | OR (95% CI) | $\chi^2$ ; *p* |
| --- | --- | --- | --- | --- |
| **Sex** |  |  |  |  |
| Male | 161 (69.4) | 71 (30.6) | 3.91 (2.62 – 5.84) | 46.193 ; < 0.001 |
| Female | 73 (36.7) | 126 (63.3) | 1 |  |
| **Age groups (years)** |  |  |  |  |
| 18-39 | 98 (50.0) | 98 (50.0) | 0.72 (0.49 – 1.06) | 2.669 ; 0.102 |
| 40-59 | 101 (62.3) | 61 (37.7) | 1.69 (1.13 – 2.52) | 6.274 ; 0.009 |
| ≥ 60 | 35 (47.9) | 38 (52.1) | 0.73 (0.44 – 1.21) | 1.427 ; 0.232 |
| **Level of education** |  |  |  |  |
| None/primary | 94 (51.9) | 87 (48.1) | 0.84 (0.57 -1.24) | 0.698 ; 0.403 |
| Secondary/higher | 140 (56.0) | 110 (44.0) | 1 |  |
| **Main occupation** |  |  |  |  |
| Farmer/Breeder | 158 (59.4) | 108 (40.6) | 1.71 (1.15 – 2.53) | 7.300 ; < 0.007 |
| Salesperson | 11 (52.4) | 10 (47.6) | 0.92 ( 0.38 – 2.22) | 0.033 ; 0.857 |
| Housewife | 21 (34.4) | 40 (65.6) | 0.38 (0.22 – 0.68) | 11.30 ; < 0.001 |
| Student | 6 (27.3) | 16 (72.7) | 0.29 (0.11 – 0.77) | 6.820 ; < 0.009 |
| Technician | 24 (72.7) | 9 (27.3) | 2.38 (1.08 – 5.26) | 4.894 ; 0.027 |
| Unemployed/Retired | 3 (30.0) | 7 (70.0) | 0.35 (0.09 – 1.38) | 2.43 ; 0.119 |
| **Community** |  |  |  |  |
| Baloua | 47 (40.9) | 68 (59.1) | 0.477 (0.30 – 0.73) | 11.388 ; < 0.001 |
| Carrière | 91 (69.5) | 40 (30.5) | 2.498 (1.61 – 3.86) | 17.461 ; < 0.001 |
| Mock-Centre | 41 (39.8) | 62 (60.2) | 0.463 (0.2 – 0.72) | 11.446 ; < 0.001 |
| Mock-Sud | 55 (67.1) | 27 (32.9) | 1.93 (1.16 – 3.20) | 6.666 ; < 0.001 |

Educational level did not appear to influence alcohol abuse (χ² = 0.698; *p* = 0.403). In contrast, occupational status had a significant impact: farmers and livestock breeders were 1.7 times more likely to experience alcohol abuse (χ² = 7.300; *p* = 0.007), and technicians had a 2.3-fold higher risk (χ² = 4.894; *p* = 0.027). Conversely, housewives showed an approximately 60% lower risk (χ² = 11.300; *p* < 0.001), while students showed a reduction in risk of alcohol abuse exceeding 70% (χ² = 6.820; *p* = 0.009) (Table 12).

Moreover, the community of residence was significantly associated with alcohol abuse. Participants living in Carrière and Mock-Sud had 2.5-fold (χ² = 17.461; *p* < 0.001) and 1.9-fold (χ² = 6.666; *p* < 0.001) higher risks, respectively, compared to residents of other communities. Conversely, those residing in Baloua and Mock-Centre showed more than a 50% reduction in risk (Baloua: χ² = 11.388; *p* < 0.001; Mock-Centre: χ² = 11.446; *p* < 0.001) (Table 12).

#### Consumption of alcoholic beverage types and risk of alcohol abuse

All types of alcoholic beverages analyzed were significantly associated with alcohol abuse, indicating that regular consumption, regardless of the beverage type, constitutes a high-risk behavior. Nonetheless, certain beverages showed a strong association with alcohol abuse. Adulterated whisky exhibited the highest association, increasing the risk of abuse by 37.9 times (χ² = 32.070; *p* < 0.001), followed by beer and palm wine, which raised the risk by 9.1-fold (χ² = 87.745; *p* < 0.001) and 5.7-fold (χ² = 63.812; *p* < 0.001), respectively. The traditional beverage “cha” was also linked to alcohol abuse, although to a lesser degree, with a 2.3-fold increase in risk (χ² = 6.406; *p* = 0.011). Moreover, regular consumption of wine, spirits, and “odontol” was consistently associated with abuse, as all regular consumers were affected; however, the small sample size for these beverages limits the strength of this finding (Table 13).

**Table 13.** Association between the consumption of alcoholic beverages and alcohol abuse.

| Variables | Alcohol abuse<br>N=185 ; n (%) | Non-abuse of alcohol<br>N=234 ; n (%) | OR (95% CI) | $\chi^2$ ; *p* |
| --- | --- | --- | --- | --- |
| **Adulterated whisky** |  |  |  |  |
| Regularly | 40 (97.6) | 1 (2.4) | 37.93 (5.16 – 278.80) | 32.070 ; <0.001 |
| Irregularly | 194 (51.3) | 184 (48.7) | 1 |  |
| **Palm wine** |  |  |  |  |
| Regularly | 136 (79.1) | 36 (20.9) | 5.74 (3.67 – 8.98) | 63.812 ; <0.001 |
| Irregularly | 98 (39.7) | 149 (60.3) | 1 |  |
| **Beer** |  |  |  |  |
| Regularly | 135 (84.9) | 24 (15.1) | 9.14 (5.54 – 15.10) | 87.745 ; <0.001 |
| Irregularly | 99 (38.1) | 161 (61.9) | 1 |  |
| **“Cha”** |  |  |  |  |
| Regularly | 35 (72.9) | 13 (27.1) | 2.32 (1.19 – 4.54) | 6.406 ; 0.011 |
| Irregularly | 199 (53.6) | 172 (46.4) | 1 |  |
| **Wine** |  |  |  |  |
| Regularly | 8 (100.0) | 0 (0.0) | 1.81 (1.66 – 1.98) | 6.448 ; 0.011 |
| Irregularly | 226 (55.0) | 185 (45.0) | 1 |  |
| **Spirits** |  |  |  |  |
| Regularly | 9 (100.0) | 0 (0.0) | 1.82 (1.66 – 1.98) | 7.272 ; 0.007 |
| Irregularly | 225 (54.9) | 185 (45.1) | 1 |  |
| **“Odontol”** |  |  |  |  |
| Regularly | 12 (100.0) | 0 (0.0) | 1.83 (1.67 – 2.00) | 9.767 ; 0.002 |
| Irregularly | 222 (54.5) | 185 (45.5) | 1 |  |

Palm wine, “Cha”, and “Odontol” are traditional beverages. “Cha” is a maize turbid beer, and “Odontol” is made from palm wine, sugar, and a tree bark.

#### Drinking motivations associated with alcohol abuse

Analysis of drinking motivations revealed that several reasons were strongly linked to alcohol abuse. Participants who reported drinking to “enjoy their evening” or to “make social activities more fun” had a 4.3-fold (χ² = 50.683; *p* < 0.001) and 4.5-fold (χ² = 53.527; *p* < 0.001) higher risk of alcohol abuse, respectively. Motivations such as “fitting in with peers” and “seeking appreciation” were even more pronounced, increasing the risk by 5.3-fold (χ² = 33.138; *p* < 0.001) and 6.4-fold (χ² = 14.905; *p* < 0.001). Similarly, drinking to “cope with nervousness or depression,” to “lift mood when feeling low,” or to “forget worries” significantly raised the likelihood of abuse, with risks increased by 3.9-fold (χ² = 9.798; *p* = 0.002), 10.3-fold (χ² = 21.383; *p* < 0.001), and 5.7-fold (χ² = 12.447; *p* < 0.001), respectively. In contrast, drinking for the “pleasure of drunkenness” was not significantly associated with alcohol abuse (χ² = 2.177; *p* = 0.140) (Table 14).

**Table 14.** Association between drinking motivations and alcohol abuse.

| Variables | Alcohol abuse<br>N=185 ; n (%) | Non-abuse of alcohol<br>N=234 ; n (%) | OR (95% CI) | $\chi^2$ ; $p$ |
| --- | --- | --- | --- | --- |
| Enjoying the evening |  |  |  |  |
| Regularly | 159 (72.3) | 61 (21.7) | 4.31 (2.85 – 6.50) | 50.683 ; <0.001 |
| Irregularly | 75 (37.7) | 124 (62.3) | 1 |  |
| Making social activities more fun |  |  |  |  |
| Regularly | 160 (72.7) | 60 (27.3) | 4.50 (2.98 – 6.80) | 53.527 ; <0.001 |
| Irregularly | 74 (37.2) | 125 (62.8) | 1 |  |
| Fitting in with peers |  |  |  |  |
| Regularly | 71 (83.5) | 14 (16.5) | 5.32 (2.88 – 9.81) | 33.138 ; <0.001 |
| Irregularly | 163 (48.8) | 171 (51.2) | 1 |  |
| Seeking appreciation |  |  |  |  |
| Regularly | 29 (87.9) | 4 (12.1) | 6.40 (2.20 – 18.55) | 14.905 ; <0.001 |
| Irregularly | 205 (53.1) | 181 (46.9) | 1 |  |
| Coping with nervousness or depression |  |  |  |  |
| Regularly | 27 (81.8) | 6 (18.2) | 3.89 (1.57 – 9.63) | 9.798 ; 0.002 |
| Irregularly | 207 (53.6) | 179 (46.4) | 1 |  |
| Lifting mood when feeling low |  |  |  |  |
| Regularly | 34 (91.9) | 3 (8.1) | 10.31 (3.11 – 34.15) | 21.383 ; <0.001 |
| Irregularly | 200 (52.4) | 182 (47.6) | 1 |  |
| Forgetting worries |  |  |  |  |
| Regularly | 26 (86.7) | 4 (13.3) | 5.65 (1.93 – 16.51) | 12.447 ; <0.001 |
| Irregularly | 208 (53.5) | 181 (46.5) | 1 |  |
| Pleasure of drunkenness |  |  |  |  |
| Regularly | 15 (71.4) | 6 (28.6) | 2.043 (0.77 – 5.37) | 2.177 ; 0.140 |
| Irregularly | 219 (55.0) | 179 (45.0) | 1 |  |

The grouped analysis of drinking motivations highlights the dominant influence of psychological, social, and conformity factors in driving alcohol abuse. Coping motives, related to managing stress or regulating negative emotions, had the strongest effect, increasing the risk of abuse by 6.28 times (χ² = 37.013; *p* < 0.001). Conformity motives followed, as drinking to respond to social pressure or a desire for acceptance raised the risk of abuse by 5.90 times (χ² = 45.642; *p* < 0.001). Social motives, involving drinking to enhance or facilitate social interactions, were also significant, multiplying the risk of abuse by 4.15 (χ² = 47.543; *p* < 0.001). In contrast, enhancement motives, oriented toward seeking positive sensations or amplifying pleasure, were not significantly associated with alcohol abuse (χ² = 2.177; *p* = 0.140) (Table 15).

**Table 15.** Association between types of motivation and alcohol abuse.

| Variables | Alcohol abuse<br>N=185 ; n (%) | Non-abuse of alcohol<br>N=234 ; n (%) | OR (95% CI) | $\chi^2$ ; $p$ |
| --- | --- | --- | --- | --- |
| **Conformity motives** |  |  |  |  |
| Regularly | 91 (83.5) | 18 (16.5) | 5.90 (3.39 – 10.26) | 45.642 ; < 0.001 |
| Irregularly | 143 (46.1) | 167 (53.9) | 1 |  |
| **Social motives** |  |  |  |  |
| Regularly | 174 (69.6) | 76 (30.4) | 4.15 (2.74 – 6.29) | 47.543 ; < 0.001 |
| Irregularly | 60 (35.5) | 109 (64.5) | 1 |  |
| **Coping motives** |  |  |  |  |
| Regularly | 71 (85.5) | 12 (14.5) | 6.28 (3.28 – 12.00) | 37.013 ; < 0.001 |
| Irregularly | 163 (48.5) | 173 (51.3) | 1 |  |
| **Enhancement motives** |  |  |  |  |
| Regularly | 15 (71.4) | 6 (28.6) | 2.04 (0.77 – 5.37) | 2.177 ; 0.140 |
| Irregularly | 219 (55.0) | 179 (45.0) | 1 |  |

Motives were classified according to Cooper's four-factor model

#### Multivariate analysis of alcohol abuse and its associated factors

Multivariate analysis revealed several factors independently associated with alcohol abuse. Among sociodemographic characteristics, being male was a significant predictor, with a 2.4-fold increased risk of abuse (AOR = 2.48; *p* = 0.002). Conversely, living in Mock-Centre had a significantly lower risk, approximately 80% less than other communities (AOR = 0.22; *p* < 0.001), while no significant associations were observed for other localities after adjustment.

Regarding drinking habits, the strongest determinant of alcohol abuse was the regular consumption of adulterated whisky, which increased the risk 26-fold (AOR = = 26.84; *p* = 0.005), followed by beer consumption (AOR = 5.81; *p* < 0.001). Among motivational factors, conformity motives had a pronounced influence, raising the likelihood of abuse by 4.8 times (AOR = 4.80; *p* < 0.001). Coping motives were also significantly associated, tripling the risk (AOR = 3.05; *p* = 0.005), while social motives contributed to a lesser extent, increasing the risk by 2.3-fold (AOR = 2.33; *p* = 0.002) (Table 16).

**Table 16.** Factors independently associated with alcohol abuse.

| Variables | Crude odds ratio |  |  | Adjusted odds ratio |  |
| --- | --- | --- | --- | --- | --- |
|  | Alcohol abuse<br>n (%) | COR (95% CI) | *p* | AOR (95% CI) | *p* |
| **Sex** |  |  |  |  |  |
| Male | 161 (69.4) | 2.48 (1.39 – 4.44) | 0.002 | 2.48 (1.39 – 4.41) | 0.002 |
| **Age groups (years)** |  |  |  |  |  |
| 40-59 years | 101 (62.3) | 1.70 (0.95 – 3.04) | 0.074 | 1.70 (0.96 – 3.03) | 0.068 |
| **Main occupation** |  |  |  |  |  |
| Farmer/Breeder | 158 (59.4) | 0.76 (0.32 – 1.81) | 0.545 | 0.78 (0.37 – 1.64) | 0.517 |
| Housewife | 21 (34.4) | 1.27 (0.44 – 3.63) | 0.652 | 1.30 (0.49 – 3.41) | 0.594 |
| Student | 6 (27.3) | 0.94 (0.24 – 3.63) | 0.935 | / | / |
| Technician | 24 (72.7) | 1.60 (0.43 – 5.89) | 0.473 | 1.65 (0.49 – 5.58) | 0.415 |
| **Community** |  |  |  |  |  |
| Baloua | 47 (40.9) | 0.59 (0.26 – 1.31) | 0.197 | 0.58 (0.26 – 1.29) | 0.187 |
| Carrière | 91 (69.5) | 0.90 (0.41 – 1.98) | 0.804 | 0.90 (0.41 – 1.98) | 0.803 |
| Mock-Centre | 41 (39.8) | 0.22 (0.09 – 0.51) | < 0.001 | 0.22 (0.09 – 0.51) | < 0.001 |
| **Adulterated whisky** |  |  |  |  |  |
| Regularly | 40 (97.6) | 26.84 (2.76 – 261.17) | 0.005 | 26.84 (2.77 – 230.20) | 0.005 |
| **Palm wine** |  |  |  |  |  |
| Regularly | 136 (79.1) | 1.54 (0.82 – 2.90) | 0.174 | 1.56 (0.84 – 2.91) | 0.159 |
| **Beer** |  |  |  |  |  |
| Regularly | 135 (84.9) | 5.81 (3.05 – 11.90) | < 0.001 | 5.81 (3.04 – 11.07) | < 0.001 |
| **“Cha”** |  |  |  |  |  |
| Regularly | 35 (72.9) | 1.10 (0.39 – 3.07) | 0.851 | / | / |
| **Conformity motives** |  |  |  |  |  |
| Regularly | 91 (83.5) | 4.82 (2.40 – 9.68) | < 0.001 | 4.80 (2.39 – 9.63) | < 0.001 |
| **Social motives** |  |  |  |  |  |
| Regularly | 174 (69.6) | 2.33 (1.35 – 4.04) | 0.002 | 2.33 (1.35 – 4.03) | 0.002 |
| **Coping motives** |  |  |  |  |  |
| Regularly | 71 (85.5) | 3.06 (1.39 – 6.73) | 0.005 | 3.05 (1.39 – 6.71) | 0.005 |

“Cha”, a traditional beverage, is a maize turbid beer

## Discussion

This study documented, for the first time in a semi-rural area of Cameroon, the prevalence of comorbidity between *Schistosoma mansoni* infection and alcohol abuse. Through this original approach, it highlights an interaction that remains little explored in the literature. It provides novel insights into the epidemiological and behavioral dynamics of this dual public health concern.

The prevalence of schistosomiasis among adults in Makenene was 34.6%, indicating a moderate endemicity at the study site according to WHO thresholds [1]. This infection rate is relatively low compared with the prevalences of 82% and 41% reported in the same locality among school-aged children in 1985 [16] and 2010 [7], respectively. Even though mass drug administration of praziquantel to school-aged children had resulted in substantial declines in prevalence across several endemic foci in Cameroon, *S. mansoni* infection is still considerable among the adult population who are not targeted by praziquantel chemotherapy [6–7].

Analysis of *S. mansoni* infection prevalence across the four surveyed communities revealed rates of 23.3%, 28.0%, 33.6%, and 50.4% in Mock Centre, Mock Sud, Carrière, and Baloua, respectively, underscoring heterogeneous transmission dynamics along the same river system (Mock River). Although all communities exhibited active endemicity, these variations highlight the influence of community-specific risk factors. The moderate prevalences recorded in Mock Centre, Mock Sud, and Carrière suggest regular contact with contaminated waters without necessarily reaching conditions conducive to hyperendemic transmission [1]. The river serves as the primary conduit for both parasite eggs and the intermediate snail host (*Biomphalaria* spp.), thereby maintaining the persistence of infection in these areas [14–15]. Community disparities could be explained by several modulatory factors, including the intensity of fecal contamination [24], the distribution and density of vector snails [14–15], as well as human behaviors and aquatic activities that likely differ between communities, thereby shaping exposure risk [27]. The particularly high endemicity observed in Baloua, with a prevalence of 50.4%, may be attributed to a combination of ecological and socio-economic conditions unique to this community. Its swampy soils provide an ideal habitat for the intermediate snail hosts of *S. mansoni*. A high density of *Biomphalaria pfeifferi*, the intermediate host of *S. mansoni*, correlated with a considerable cercarial shedding rate has been previously reported in Baloua [15]. Moreover, the swampy soil and irrigated farmlands of the Baloua neighborhood, sustained by water from the Mock River, enable off-season farming, a common agricultural practice that serves as a major anthropogenic driver of hyperendemicity by increasing human–water contact.

Beyond community-level differences, sociodemographic analysis indicates that sex was not a significant factor of variation in our study population, aligning with findings from other investigations conducted in endemic African contexts [28–30] and even within the same study site among school-aged children [17]. This trend may be explained by the fact that men and women, although engaged in distinct daily tasks, ultimately share a comparable level of exposure to contaminated water sources. Women are frequently exposed during household chores such as laundry, dishwashing, and water collection, and occasionally through agricultural activities, whereas men are more frequently exposed through farming, fishing, or watering livestock. The reliance on natural water bodies thus represents both a daily necessity and a culturally embedded practice, minimizing gender-based differences and accounting for the relatively similar prevalence observed between sexes.

In contrast, age emerged as a high-risk factor for *S. mansoni* infection, with a clear declining trend in infection rates as age increased. This observation may be explained by reduced exposure over time. Young adults (18–39 years) are more actively involved in high-risk activities, either for income-generating purposes or recreational pursuits. Older individuals, on the other hand, often demonstrate more cautious behaviors, reinforced by progressive physical limitations, such as choosing safer water sources. In addition, repeated exposures to the parasite over a lifetime may lead to the development of partial acquired immunity, which can reduce parasite burden or decrease the risk of reinfection in adulthood. Similar patterns have been reported in several studies from sub-Saharan Africa, where prevalence declines after peaking in adolescence/early adulthood [31–33]. Occupational status also emerged as a key determinant of risk to *S. mansoni* infection, with farmers and breeders being particularly exposed due to their frequent and prolonged contact with contaminated water sources. As highlighted by several authors, irrigated agricultural practices substantially contribute to the persistence of schistosomiasis in rural areas of sub-Saharan Africa [25, 27, 33–35].

The present study also highlighted problematic alcohol consumption, with an overall prevalence of 54.3% for alcohol abuse, which combines harmful use of alcohol and alcohol dependence. This particularly high level fits within a concerning national context. According to the WHO, Cameroon ranks among the African countries with the highest per capita alcohol consumption, with rural areas appearing especially vulnerable due to abundant and poorly regulated availability [12]. In Makenene, the proliferation of drinking establishments, the wide availability of low-cost traditional drinks (palm wine, “cha,” and “odontol”), the scarcity of healthy recreational infrastructures and activities, as well as the absence of effective regulation on minimum drinking age and sales hours, all represent structural drivers of excessive alcohol consumption [40]. Illustratively, 16% of the participants in this study reported having started to drink alcoholic beverages more regularly between the ages of 10 and 15, and 40% between 16 and 20 years old. Given that the legal drinking age in Cameroon is 18 years, these data highlight the poor prevention and regulatory policies of abusive alcohol consumption. These findings are consistent with observations by Rabotata *et al*. [36] and Martín-Turrero *et al.* [37], who emphasize the major role of multiple points of sale and the financial accessibility of alcoholic beverages in driving alcohol abuse in resource-limited contexts.

Disparities in alcohol consumption patterns by sex are a globally observed phenomenon, with men tending to consume larger quantities and at higher frequencies [12, 38]. The findings of this study align with this trend and are consistent with WHO estimates for 2019 regarding sex-specific levels of alcohol consumption in Cameroon [12]. This pattern can be attributed to a combination of physiological, sociocultural, and psychological factors: men’s stronger biological sensitivity to the harmful effects of alcohol may influence consumption behaviors [39, 40]; social pressure to drink is more pronounced in public and economic spheres; and cultural norms often valorize alcohol as a marker of masculinity and conviviality [41, 42]. Although women are less affected, they are not exempt from risk, particularly in rural contexts where collective celebrations (funerals, weddings, traditional festivals) foster consumption. However, the persistent stigma attached to female drinking may account for their lower apparent involvement [42–45].

Age also showed a notable influence, with middle-aged adults (40–59 years) presenting a higher likelihood of alcohol abuse. This trend, observed in other African contexts as well [46], may be explained by cumulative exposure over time and the deeper integration of drinking habits into social and professional routines. Furthermore, the data suggest that occupational status constitutes an indirect factor of exposure, with farmers being the most represented among regular drinkers. This tendency may be linked to the strenuous nature of agricultural work, which can drive some individuals to use alcohol as a means of relieving fatigue, enhancing sociability, or managing stress [38, 40]. The sociocultural context also plays a central role. This study revealed that social and conformity motives were strongly associated with alcohol abuse, confirming that drinking is often perceived less as an individual need than as a group norm [38, 40, 47].

A joint analysis of the data revealed a notable comorbidity between *Schistosoma mansoni* infection and alcohol abuse, with a prevalence of 17.5%. This reflects a concerning public health situation [1]. The coexistence of these two conditions can be explained by the persistent schistosomiasis endemicity in these communities alongside high levels of alcohol consumption, particularly in its harmful forms. This dual burden further weakens already vulnerable populations, in line with the observations of Teixeira *et al*. [48], Pillai *et al*. [49], and Tharmalingam *et al.* [50] on the immunosuppressive effects of alcohol that may facilitate parasitic infections, as well as Belete *et al.* [51], Goma *et al.* [52], and Turyasiima *et al.* [53] on harmful alcohol consumption in sub-Saharan Africa.

Residence emerged as a determinant factor: certain communities, such as Baloua and Carrière showed higher levels of comorbidity, reflecting both persistent parasite transmission and cultural habits that encourage the consumption of alcoholic beverages. These inter-community disparities echo the “persistent foci” described in several studies [54–55], where schistosomiasis remains endemic despite control campaigns, while also aligning with the analyses of Sudhinaraset *et al*. [56], who emphasize the influence of social and cultural norms on alcohol consumption. Moreover, the results of this study highlight specific behavioral practices that reinforce the risk of comorbidity. Regularly performing household tasks at transmission sites significantly increased the risk of *S. mansoni* infection, when frequent consumption of certain drinks, such as adulterated whisky, beer, and palm wine, was strongly associated with alcohol abuse. Adulterated whisky exhibited an exceptionally high odds ratio, highlighting its strong addictive potential linked to its high alcohol content (approximately 43% for varieties marketed in Cameroon), and contextual factors such as its very low cost and wide availability, even in small retail shops. This combination enhances accessibility and exacerbates the risk of overconsumption within communities. These findings are consistent with those of Ferreira-Borges *et al.* [57] and Staton *et al.* [58], who identify inexpensive local alcoholic beverages as major drivers of alcohol abuse.

Motivations underlying alcohol consumption constitute another crucial factor in understanding this comorbidity. Social and conformity-related aspects, such as seeking conviviality and enhanced social integration, were strongly associated with alcohol abuse. These findings, consistent with several other studies [38, 41, 47], corroborate the motivational models of Cooper [20] and Kuntsche *et al.*, which classify social and psychological motives as the main predictors of excessive consumption [59]. The presence of these determinants suggests that schistosomiasis-alcoholism comorbidity is not merely the sum of two distinct problems but reflects shared social and psychological vulnerabilities that amplify risks.

Some limitations should be considered when interpreting our findings. First, the use of different diagnostic approaches for *S. mansoni* infection, POC-CCA for a minority of participants and Kato-Katz for the majority, may have introduced variability in prevalence estimates. This methodological heterogeneity could have resulted in an underestimation of *S. mansoni* infections. In addition, assessing alcohol consumption levels and risk factors for both conditions relied exclusively on self-reported data, which is subject to recall and social desirability bias, potentially underestimating the true extent of these factors and their contribution to the observed dynamics.

## Conclusion

This study provides, for the first time, documented evidence of comorbidity between *Schistosoma mansoni* infection and alcohol abuse in a semi-rural area of Cameroon. The findings highlight a moderate prevalence of schistosomiasis among adults, coupled with problematic alcohol use affecting more than half of the participants. The analysis of sociodemographic and behavioral factors reveals that regular consumption of high-alcohol-content and adulterated beverages, along with factors associated with agricultural activities and frequent human-contaminated water sources, exacerbates this dual health burden.

The identification of this interaction underscores the need to integrate alcohol abuse prevention into schistosomiasis control strategies to enhance the effectiveness of public health interventions. Combined approaches, including chemoprevention, reduction of environmental risk factors, and the promotion of responsible drinking behaviors, are essential to advance toward the elimination targets for schistosomiasis while mitigating the impact of alcohol abuse on population health.

## Data Availability

All relevant data are within the manuscript

## Declarations

### Consent for publication

Not applicable

## Acknowledgments

The authors express their sincere gratitude to the Ndikinimeki Health District Chief, the Chief doctor of the Makenene Sub-district Health Center, and its personnel for their institutional support. We also extend our gratitude to the community health workers of Makenene for their role in mobilization and sensitization, as well as to all the participants who took part in this study.

Special acknowledgment is extended to the laboratory technician Ms Lieheu Germaine for her essential contribution to parasitological examinations, and to Dr. Elong Jules for his support on data analysis.

## Availability of data and materials

All relevant data are within the manuscript

## Competing interests

The authors declare that they have no competing interests

## Funding

ESY was supported by the Children’s Investment Fund Foundation (CIFF) as part of the RSTMH Small Grants Program 2021, under the supervision of HBJ. The funder had no role in study design, data collection and analysis, decision to publish, or preparation of the manuscript.

## Author contributions

**Conceptualization**: Hermine B. Jatsa, Christian M. Kenfack, Ulrich M. Femoe, Emmanuelle S. Yimgoua, Louis-Albert Tchuem Tchuente

**Data curation**: Hermine B. Jatsa, Emmanuelle S. Yimgoua

**Formal analysis**: Hermine B. Jatsa, Emmanuelle S. Yimgoua

**Investigation:** Emmanuelle S. Yimgoua, Christian M. Kenfack, Nestor G. Feussom, Joseph B. Fassi-Kadji, Emilienne T. Nkondo, Ulrich M. Femoe, Hermine B. Jatsa

**Methodology**: Hermine B. Jatsa, Emmanuelle S. Yimgoua, Christian M. Kenfack, Nestor G. Feussom, Joseph B. Fassi-Kadji, Emilienne T. Nkondo, Ulrich M. Femoe

**Funding acquisition**: Emmanuelle S. Yimgoua

**Project administration**: Hermine B. Jatsa

**Supervision**: Hermine B. Jatsa, Louis-Albert Tchuem Tchuente

**Validation**: Hermine B. Jatsa, Louis-Albert Tchuem Tchuente

**Writing – original draft**: Emmanuelle S. Yimgoua, Hermine B. Jatsa

**Writing – review & editing:** Hermine B. Jatsa, Louis-Albert Tchuem Tchuente

## Notes

### Competing Interest Statement

The authors have declared no competing interest.

### Funding Statement

Yes

### Author Declarations

This study was approved by the Regional Committee of Research Ethics for Human Health of the Centre region, under the authority of the Ministry of Public Health of Cameroon (CE N° 213/CRERSHC/2022). Informed consent was obtained from all participants prior to inclusion in the study.

